# New RDEB intermediate variant with in-frame partial exon skipping in FN III-like domain of type VII collagen

**DOI:** 10.1101/2022.09.02.22278356

**Authors:** N.A. Evtushenko, A.A. Kubanov, A.A. Martynova, N.V. Kondratyev, A.K. Beilin, A.E. Karamova, E.S. Monchakovskaya, К.A. Azimov, M.A. Nefedova, N.G. Bozhanova, E.V. Zaklyazminskaya, N.G. Gurskaya

**Affiliations:** Pirogov Russian National Research Medical University: Moscow, Russia; State Research Center of Dermatovenerology and Cosmetology, Russian Ministry of Health: Moscow, Russia; Mental Health Research Center: Moscow, Russia; Vanderbilt University: Nashville, TN, USA; Petrovsky National Research Centre of Surgery, Russia

## Abstract

Recessive Dystrophic Epidermolysis Bullosa (RDEB) is a debilitating genodermatosis caused by pathogenic mutations in the *COL7A1* gene, which induce absence or reduction in the number of anchoring fibrils. The severity of RDEB depends on the mutation type and localization, but many aspects of this dependence remain to be elucidated. Here, we report a novel variant of RDEB Intermediate in two unrelated patients. Their disease manifestation includes early skin and oral mucosa blistering and is associated with localized atrophic scarring. According to the exome and Sanger sequencing results, both investigated Probands are the carriers of complex heterozygosity in the *COL7A1* gene with the same deletion in intron 19 of the *COL7A1* gene. RT-PCR followed by sequence analysis revealed skipping of the part of exon19, as well as the rescue of the open reading frame (ORF) of *COL7A1* in both Probands. We hypothesize that the mutation in the acceptor splice site leads to the activation of the cryptic donor splice site, resulting in the truncated but partially functional protein and the milder phenotype of intermediate RDEB. This rare type of mutation expands our understanding of RDEB etiology and invites further investigation.

## Main text

Recessive Dystrophic Epidermolysis Bullosa (RDEB) is a genodermatosis characterized by blister formation at the level of sublamina densa. It is an incurable disease caused predominantly by mutations in collagen type VII alpha 1 chain encoding gene (*COL7A1*) (Eichstadt et al., 2019). Although RDEB is a disabling disease, the severity of symptoms varies greatly and largely depends on the type and localization of mutations, as well as the number of yet unknown congenital factors (Dang and Murrell, 2008; Odorisio et al., 2014). *COL7A1* gene contains 118 exons and is enriched in highly repetitive structures.

Collagen VII (COLVII) α1 protein, encoded by the *COL7A1* gene, consists of a central helical domain that is surrounded by two terminal noncollagenous domains (NC-1 and NC-2). The amino-terminal NC1 domain contains two von Willebrand A factor domains split by nine fibronectin III (FN-III)-like domains. Triple-helical domain consists of the canonical Gly-X-Y sequence. NC-2 does not contain repeating structures, but the structure is essential for dimerization (Christiano et al., 1994).

Most exons of *COL7A1* are in-frame, and natural exon-skipping was described for the gene (Bremer et al., 2019). It was demonstrated that the deletion of the repeating-motif exons does not perturb protein function (Bornert et al., 2016; Goto et al., 2006; Mencía et al., 2018). Most cases of exon-skipping are attributed to collagenous domains (Turczynski et al., 2016) with only few positioned in FN-III (Bremer et al., 2019; McGrath et al., 1999). Among the latter is the skip of the entire exon 19 that removes 49 amino acids from within the seventh FN-III. It was shown to restore the reading frame for *COL7A1* (Mellerio et al., 1999).

Here, we report two unrelated RDEB cases with a new mutation that alters the acceptor splicing site in exon 20 of *COL7A1*. Interestingly, these patients reveal only relatively mild symptoms of RDEB Intermediate.

Proband I is a 23-28-year-old male, who was admitted to the dermatology department with skin and mucous blistering. There was no familial history of EB, including his younger sibling. In the first weeks of life, the localized blistering and erosions started to occur on upper and lower limbs spreading to other skin areas and oral mucosa subsequently. At preschool age, this patient has manifested anonychia, ankyloglossia, microstomia, and partial adentia, then esophageal stricture, foot deformity, and pseudosyndactyly. At present, the physical examination revealed atrophic scarring localized on hands and shins accompanied by sporadic blisters. Surgical release of pseudosyndactyly was performed (photographic materials can be provided by the corresponding author upon reasonable request).

Proband II is a 13-18-year-old male with blistering and erosions on skin and oral mucosa, and atrophic scarring. His parents and sibling were reported to have no clinical signs of EB. Skin blistering on feet was presented at birth. In the first few months after birth hands started to be affected by the pathological process as well. Esophageal stricture, musculoskeletal contractures, pseudosyndactyly, and other extracutaneous manifestations are not presented in this patient. Clinical features of Proband II point to RDEB Intermediate form (photographic materials can be provided by the corresponding author upon reasonable request).

Immunofluorescent staining (IF) of skin paraffin sections with anti-COLVII antibodies demonstrated the reduced COLVII protein level in dermal-epidermal junction (DEJ) zone in the skin of the Proband I (Figure 1A) in comparison with the control sample of a healthy donor (Figure 1B).

**Figure 1.**
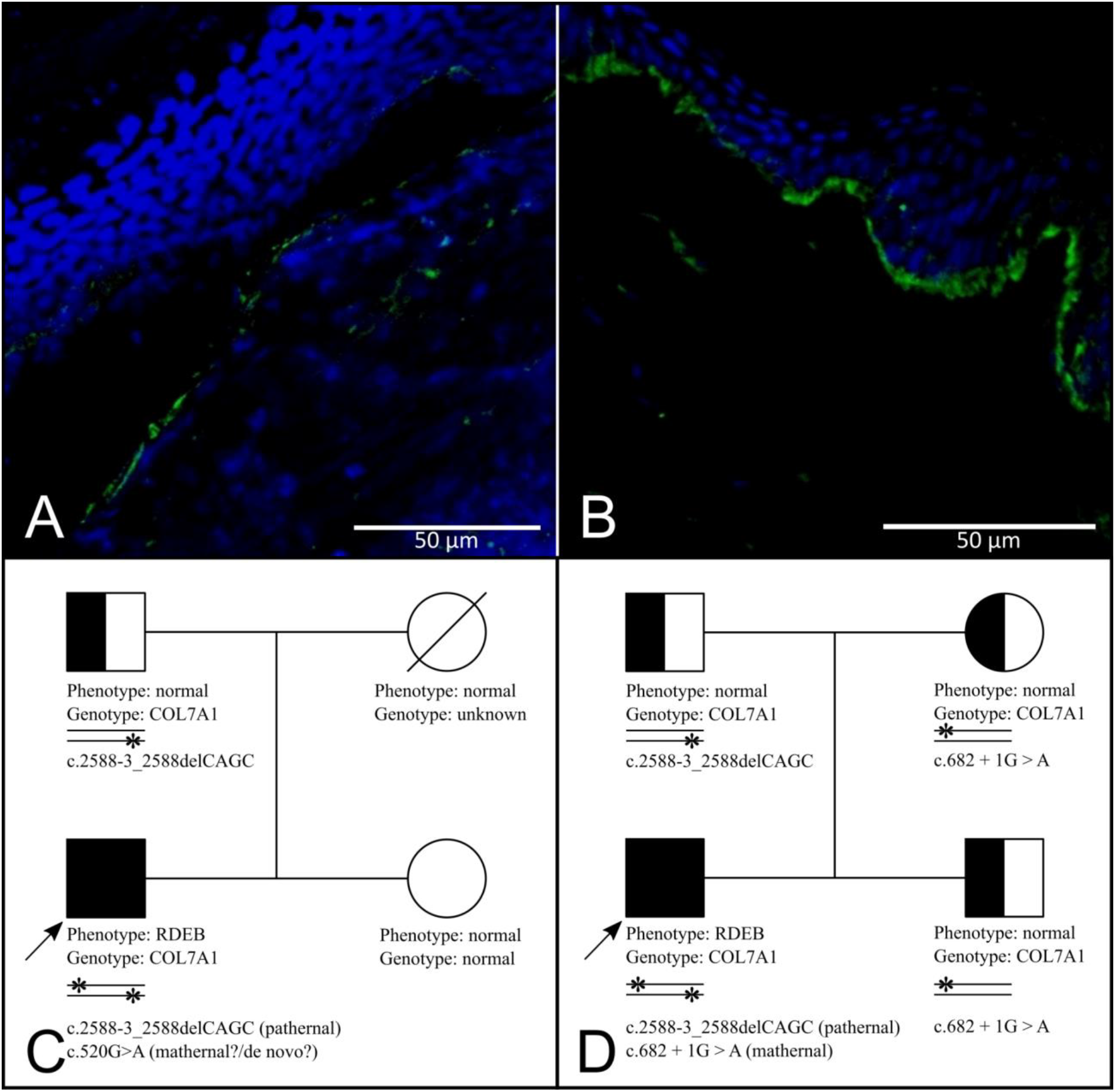
Immunostaining analysis of the skin biopsies and pedigrees of the patients. **A, B** – Immunofluorescent staining (IF) of skin paraffin sections of Proband I (**A**) and a healthy donor (**B)**. Anti-collagen VII (COLVII) rabbit (Rb) polyclonal antibody, 0.5 mg/ml, GeneTex, USA, GTX37733, diluted 1:80 and goat (Gt) anti-rabbit polyclonal antibody DyLight 488, 1 mg/ml, diluted 1:400. Stained nuclei are visible in blue channel (DAPI). **C**. Pedigree of Proband I. Sanger sequencing of the *COL7A1* gene confirms deletion in heterozygous state in the paternal sample, whereas no mutations were found in the sister’s DNA sample. **D**. Pedigree of Proband II. Sanger sequencing of the *COL7A1* gene revealed a deletion in the paternal sample and mutation c.682+1G>A in intron 5 in the maternal DNA, both in heterozygous state. Proband’s brother inherits only maternal mutation. The arrows indicate the probands. Written consents (from patients and parents of the patients) were obtained for publications of the pedigrees.

Initially, to understand the nature of pathological changes, we performed exome sequencing of the blood sample of Proband I (Novagene, USA) with approximately 23 million reads (2×150 PE). The reads were aligned to the human genome assembly (hg38) with HISAT2 and processed with a single-sample germline SNP and indels discovery pipeline according to the Broad Institute’s tutorial (Germline short variant discovery (SNPs + Indels), n.d.). The analysis yielded two pathogenic protein-altering mutations, with apparent relation to the phenotype of interest (See Supplementary Table S1 for a detailed description of Variant Calling). Two variants were found to be splice variants located inside the *COL7A1* gene, both heterozygous, hereafter referred to as c.2588-3_2588delCAGC and c.520G>A. The mutation c.2588-3_2588delCAGC is new and has not been yet reported in genetic databases, while c.520G>A is known as a rare SNP rs1245411910 in dbSNP v155 and annotated as pathogenic in Clinvar. We confirmed paternal inheritance of the deletion, as Sanger sequencing revealed c.2588-3_2588delCAGC heterozygous mutation in the healthy father of the Proband I (Figure 1C). The heterozygosity of Proband’s mutations was additionally validated by Sanger sequencing of the DNA fragment that covered both sites of mutations extending from the beginning of Exon4 to the end of Exon20 (Figure 2A,B and data not shown). Mutations of Proband I were registered in LOVD open variant database (LOVD - Leiden Open Variation Database, An Open Source DNA variation database system: Variant #0000763600, n.d.).

**Figure 2.**
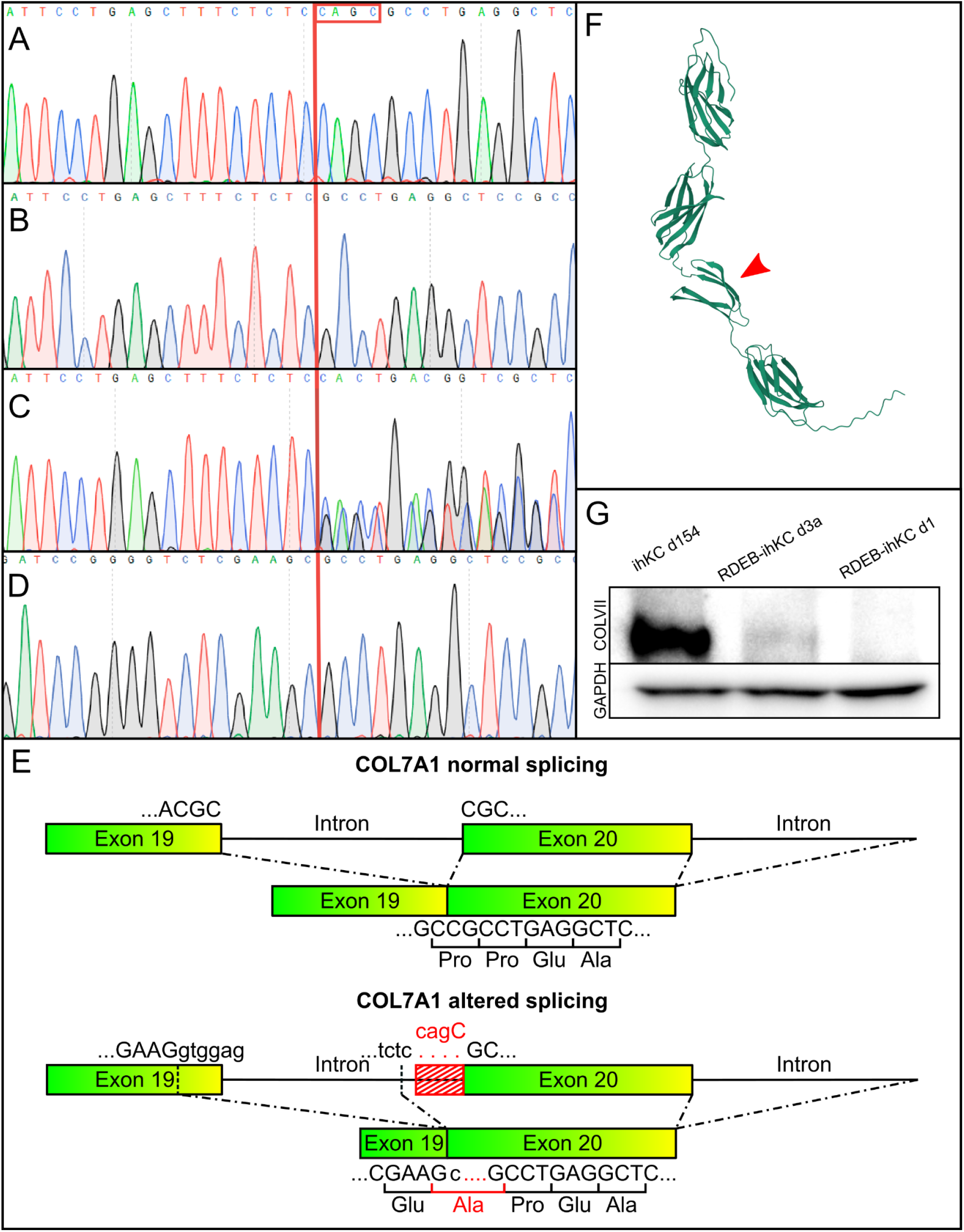
Novel mutation in the *COL7A1* gene and its impact on the splicing, COLVII protein folding and expression level. **A-C**. Sequences of the intron 19/exon 20 junction regions of the *COL7A1* gene variants. Red line denotes the area of CAGC deletion. **A**. *COL7A1* wild type allele of Proband I, the CAGC sequence is highlighted. **B**. *COL7A1* c.2588-3_2588delCAGC allele of Proband I. **C**. Heterogeneity in the *COL7A1* gene of Proband II. **D**. Sequence of exon 19-20 junction region of cDNA from c.2588-3_2588delCAGC allele. **E**. Schematics of normal splicing (top) and abnormal splicing due to CAGC deletion (bottom). Skipping of the part of exon 19 appears to be the result of activation of the internal exonic cryptic donor splice site (DSS). The position of the cryptic DSS is marked by a dashed line; deleted nucleotides are shown in red. **F**. Structure of the FN III-like domains 6-9 predicted by trRosetta. Abnormal seventh FN III-like domain is lacking two out of seven β-strands. **G**. Western Blot analysis of lysates from immortalized keratinocytes of a healthy donor (ihKC154), Proband I (RDEB-ihKC d3a), and an RDEB patient with homozygous c.425A>G (RDEB-ihKC d1), 6% PAAG, Goat antibodies against COLVII (GeneTex), Rabbit antibodies against GAPDH, Electro-chemiluminescence (ECL) detection.

We further found that Proband II also possesses compound heterozygosity. Sanger sequencing of the blood sample revealed mutations in the intron 5 (c.682+1G>A) and the same four base pairs deletion c.2588-3_2588delCAGC as in Proband I (Figure 2C). Both parents and the sibling of the patient are healthy heterozygous carriers of the mutant alleles (Figure 1D).

Both Probands possess the complex heterozygosity in the *COL7A1* gene. They share one mutation, c.2588-3_2588delCAGC, and each has an additional splicing-affecting mutation in their second allele: c.520G>A or c.682+1G>A. The mutation of the last nucleotide in exon 4, c.520G>A, disturbs the splicing of the downstream intron. It was registered in LOVD database as p.G174Arg missense mutation (Almaani et al., 2011). Our sequence data of cDNA clones revealed splicing via a new donor splice site (DSS) in the close proximity of the former DSS, ORF shift, and a premature termination codon (Figure S1). The mutation of Proband II c.682+1G>A is at the beginning of intron 5 and it also disrupts the DSS (Hovnanian et al., 1997).

In addition, we found an SNP c.2817A>G (p.Pro939=) in both Probands. This variant is known as rs1264194 and is denoted in ClinVar as benign. However, its frequency was found to be increased among Spanish RDEB patients (Cuadrado-Corrales et al., 2010).

This study aims to reveal the effect of the novel c.2588-3_c.2588delCAGC variant of the *COL7A1* gene.

We hypothesized that the mutation c.2588-3_2588delCAGC in intron 19 induces a splicing impairment since it localizes at the acceptor splice site (ASS). Exon skipping events due to ASS mutations were shown before for introns 86 and 107. In both cases, the downstream exons (87 and 108, correspondingly) were skipped (Bremer et al., 2019). The skipping of the whole exon 19 was observed previously as a result of duplication in c.2471 of exon 19 (Bremer et al., 2019; McGrath et al., 1999). To investigate the mutation effect, we performed RT-PCR followed by cDNA sequencing (Figure S2, Supplementary Table S2). Surprisingly, despite this deletion destroying a consensus splice acceptor pattern NYAG, exon 20 was fully preserved while a part of the upstream exon 19 was lost (Figure 2D&E). This exon 19 partial skipping removes 81 base pairs, which does not disturb the ORF of *COL7A1*, and leads to the single Alanine>Proline substitution at the exon 19-20 junction (p.(Gly837_Pro863delinsAla), Figure 2 D, E).

The observed partial exon skipping might be explained by the activation of cryptic splice elements. Indeed, Human Splicing Finder 3.0 (HSF3.0) (Genomnis_HSF_Pro, n.d.) revealed a rare noncanonical ASS motif TCTC at the 3’ side of the intron 19. Both HSF3.0 and Alternative Splice Site Predictor (ASSP - Prediction, n.d.) suggested the presence of a cryptic DSS c.2504-2512 in exon 19 (c.2506 position is the last in the mutant truncated exon 19) (Figure S3A). Additionally, exon splicing enhancers (ESE) were located by HSF3.0 at the beginning of exon 20 (Figure S3B).

Spicing efficiency regulation is multifaceted. Among others, exon inclusion depends on the distance between splicing sites, the strength of splice sites, the distribution of ESE and ESS, and the secondary structure of pre-mRNA (Ptok et al., 2019). According to HSF 3.0, the DSS in the middle of exon 19 is stronger than the natural DSS at the end of exon 19 (Table S3). Therefore, when the canonical ASS is deleted, the cryptic DSS might be preferred due to its strength despite the larger distance from ASS.

Although the aberrant splicing occurred with low efficiency, the produced protein might be partially functional. *COL7A1* exons 6-24 encode nine consecutive FN III-like domains. The skipped part of exon 19 removes 27 amino acid residues from the seventh FN III-like repeat. We used trRosetta (Yang et al., 2020) implemented on the Robetta server (Robetta, n.d.) to model the structure of the probands’ FN III-like domains 6-9 region. According to the obtained model, the missing amino acid residues were forming two full consecutive β-strands of the seventh domain (Figure 2F, & Supplement pdb file). We hypothesize that, despite this deletion, at least a fraction of the expressed protein might successfully fold and retain a close-to-nature structure, allowing for partial or full functionality.

To further test our hypothesis about the presence of aberrant COLVII protein in the probands’ cells we analyzed the COLVII levels in cultured cells of Proband I. We isolated keratinocytes from Proband I skin biopsy after obtaining informed consent and immortalized them according to the previously published protocol (Evtushenko et al., 2021). We investigated the amount of the COLVII protein in the keratinocytes’ lysates by Western blot analysis (Figure 2G). The analysis revealed residual amounts of COLVII in the Proband I keratinocytes. This suggests that the p.(Gly837_Pro863delinsAla) COLVII indeed is capable of forming a folded protein and might be responsible for the alleviated RDEB phenotype of probands.

In conclusion, the novel c.2588-3_2588delCAGC mutation of *COL7A1* perturbs ASS and induces a pathogenic phenotype with mild features of RDEB generalized-intermediate. We have shown that this mutation leads to the activation of the alternative splice sites and induces skipping of the part of exon 19 while preserving the reading frame. Skipping of several amino acids is potentially less deleterious than expression of a truncated protein or nonsense-mediated mRNA decay. Moreover, these data point to a region of FN III-like domains as promising for the application of exon skipping therapy to rescue pathological RDEB mutations (Turczynski et al., 2016).

## Supporting information

Fibronectin 5-9 domains structure

## Data Availability

All data produced in the present study are available within the manuscript and supplementary materials or from the corresponding author on reasonable request.

## Acknowledgments

We thank professor Anton A. Komar (Cleveland State University) for the assistance with access to Novagene and for fruitfull discussion. We thank professor Johan den Dunnen (Leiden University) for help in data curation in LOVD.

## Author Contributions

Conceptualization: NG, AK;

Data Curation: NE, NG, NB;

Formal Analysis: NE, NK, NG;

Funding Acquisition: AnAK, NG;

Investigation: NE, KA, AM, MN, NK;

Methodology: NK, AEK;

Software: NK, NB;

Supervision: AK, NG;

Figures Design: NE,AB;

Visualization: EM, AEK, AB;

Original Draft Preparation: NE, NG, NB;

Writing and Editing:NE, NG, NB

## Data availability statement

The study was conducted in accordance with the Declaration of Helsinki, and approval was obtained from the Local Research Ethics Committee of Pirogov Russian National Medical University (Protocol No. 219 of 2 June 2022). Informed consents were obtained from all patients involved in the study. Written informed consents were obtained from patients to publish their images or case history. All other relevant data supporting the key findings of this study are available within the letter and Supplementary Materials or from the corresponding author on reasonable request.

## Supplementary Materials

