## Supplementary Materials for "New RDEB intermediate variant with in-frame partial exon skipping in FN III-like domain of type VII collagen"

**Table S1. Exome sequencing data.**

**Table S2.** **The oligonucleotides sequences used for RT-PCR amplification of *COL7A1* fragments and for nested PCR amplification before sequencing.**

| Primer name | Primer sequence 5’- 3’ |
| --- | --- |
| fw Ex 16 С7 | GCCACAGGATACAGGGTTTC |
| rev Ex 22 | CACCACACGTAGTTCAATGC |
| fw Ex 17 C7 (nested) | GACTGGAGCCAGATACTGAGTAT |
| fw Ex 17-18 (nested, exon junction) | GTGAGGACTGCCCCTGAGCCTG |
| rev Ex 21 (nested) | CCTCACGCGGTACTGTGTC |
| rev Ex 22-21 (nested, exon junction) | ACGAGGTGACTCAGTGCGCGCA |
| fw 1 Ex 18 | CCCCTGAGCCTGTGGGTCGT |
| nest fw Exon 18 | GTGTCGAGGCTGCAGATCCTC |
| rev Ex 20 (nested) | GAACAGCTCCTCGCCCCCGCTAGGTTGCCAGTGCAGAAGGAAGC |
| rev Ex 22 | CACCACACGTAGTTCAATGC |

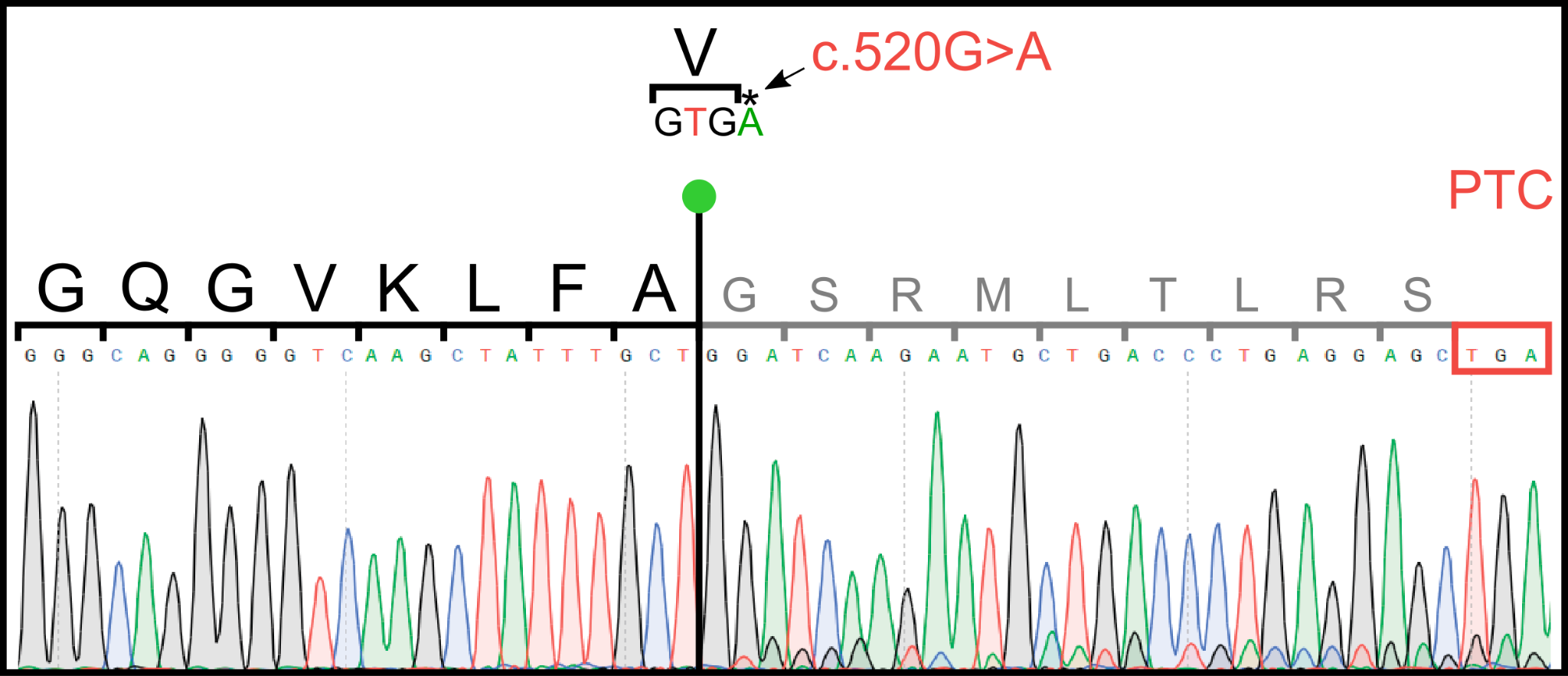

**Figure S1. Effects of the mutation of the last nucleotide in exon 4, c.520G>A, in Proband I on *COL7A1* splicing.** Shown is the sequence of the cDNA fragment at the abnormal exon 4/exon 5 junction site (denoted by the red line). A new donor splice site (DSS) causes omitting of Val173 (shown above) and the ORF shift, resulting in the premature termination codon (PTC) insertion and making the transcript suitable for nonsense-mediated mRNA decay. Black line with green pinhead denotes the new 3’ exon 4 boundary.

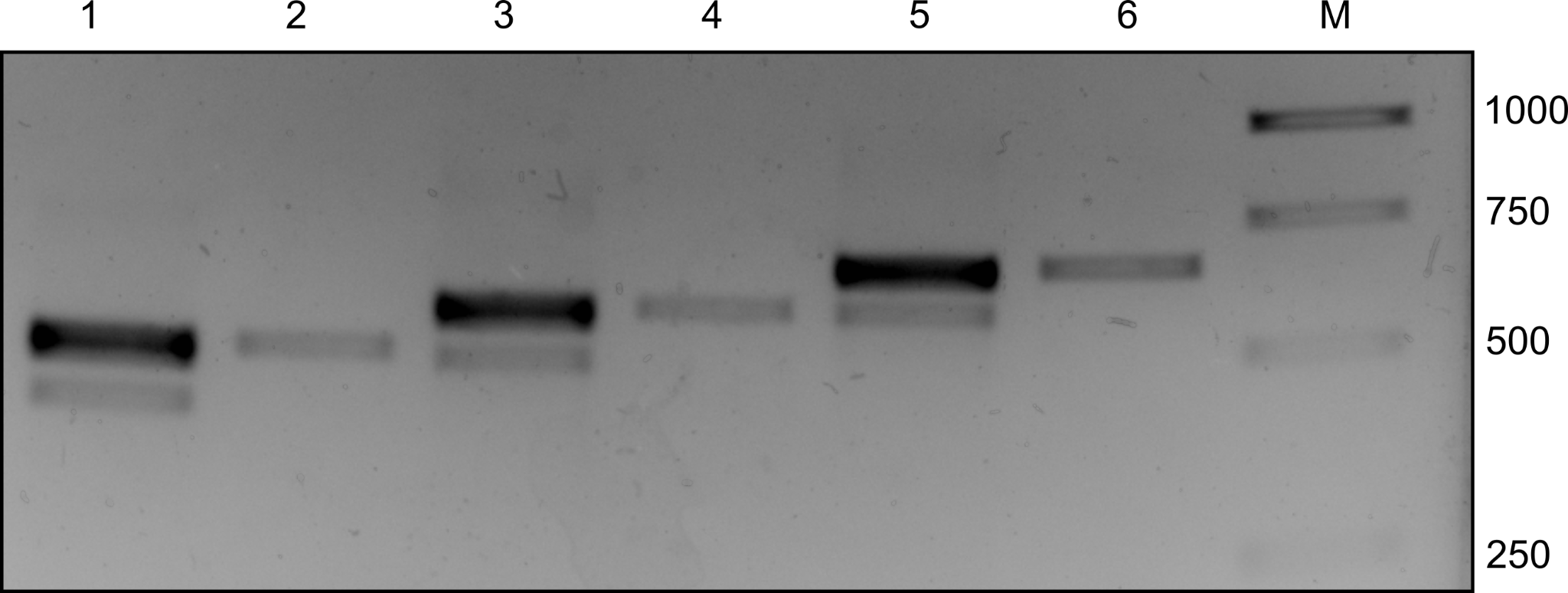

**Figure S2.** **RT-PCR analysis of *COL7A1* cDNA from Proband I and a healthy donor.** Shown are results of PCR amplification using different pairs of specific primers. Lanes 1, 3, 5 - Proband I; lanes 2,4, 6 - healthy donor. Amplification with primers Ex17-18Fw and Ex21Rv: 1, 2. Amplification with primers Ex17-18Fw and Ex21-22Rv: 3, 4. Amplification with primers Ex17Fw and Ex21-22Rv: 5, 6. M - Marker of molecular weight.

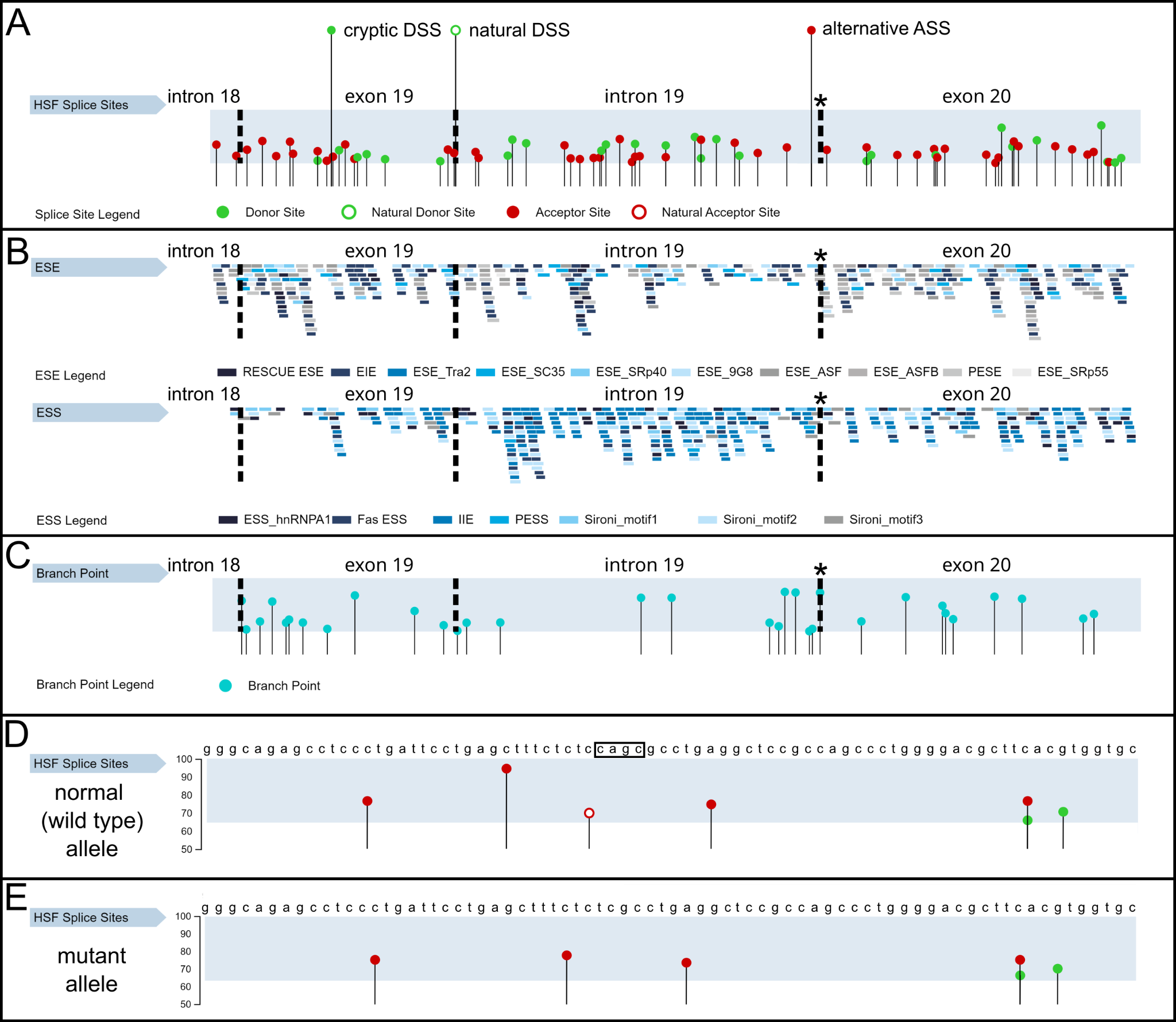

**Figure S3.** **HSF 3.0 prediction of splicing signals in *COL7A1* (Genbank accession no.NG_007065/1) fragments.**

**A.** The splice sites localization in the DNA fragment of exon 19, intron 19, and exon 20 of *COL7A1* containing deletion c.2588-3_c.2588delCAGC. Asterisk marks the location of the deletion. Green pinheads denote DSSs, red pinheads denote ASSs. Black dashed lines denote exonic boundaries. Precise values (score) of predicted splice sites are available in the Table S3. **B**. The predicted binding sites of Exonic Splice Enhancers (ESE), Exonic Splice Silencers (ESS), exonic identity element (EIE), putative exonic splicing enhancer (PESE) and silencer (PESS), found in the *COL7A1* c.2588-3_c.2588delCAGC gene. Several groups of ESE, ESS are shown that recognized by different splice factors. **C**. Branch points localization in the fragment of *COL7A1* c.2588-3_c.2588delCAGC. Several branch points are predicted in the area of the skipped part of exon 19. **D, E**. Splice sites localization in the intron 19/exon 20 region in wild type allele of *COL7A1* (**D**) and in *COL7A1* c.2588-3_c.2588delCAGC (**E**). Red pinheads denote ASSs. Red with white pinhead denotes natural ASS. Black frame denotes the CAGC motif that is deleted in the mutant allele.The difference in ASS distribution is clearly visible: the alternative ASS is formed while the natural ASS disappears in the mutant sequence (Fairbrother et al., 2004).

**Table S3. HSF Signals of Splicing of *Col7A1* exon19-exon20 fragment with deletion c.2588-3_c.2588delCAGC**

**Supplementary Methods**

Immunofluorescent (IF) staining was performed with paraffin sections according to standard protocol. We used primary rabbit polyclonal antibody to collagen VII (GeneTex, USA, GTX37733) and secondary goat anti-rabbit polyclonal antibody DyLight 488 (GeneTex, USA, GTX213110-04).

For Western Blot (WB) analysis cells were lysed in RIPA buffer and separated in 6% acrylamide gel without urea alongside a molecular weight marker (161-0377, Bio-Rad, Hercules, California, USA). Semidry transfer on nitrocellulose membrane was performed with the Trans-blot Turbo Transfer system (Bio-Rad, Hercules, U.S.). Immunostaining was performed with polyclonal Goat antibodies against NC1 peptide of type VII collagen (GeneTex, GTX89040, dilution 1:1000), and anti-GAPDH Rabbit antibodies (ab9485, dilution 1:4000). ImmPRESS® HRP Horse Anti-Goat IgG Polymer Detection Kit, peroxidase (Vector Laboratories, Burlingame, U.S., MP-7405), Clarity Western ECL Substrate (Bio-Rad, Hercules, U.S.) were used for Electro-chemiluminescence detection in the ChemiDocMP Imaging System (Bio-Rad, Hercules, U.S.).

All synthetic DNA oligonucleotides for cloning and RT-PCR were purchased from Syntol (Moscow, Russia) and Evrogen (Moscow, Russia). RT-PCR was performed by MMLV-kit (Evrogen, Moscow, Russia) according to the manufacturer’s instructions. DNA fragments were cloned into the pAL-TA (Evrogen, Moscow, Russia), and the plasmids were used for Sanger sequencing (Evrogen, Moscow, Russia).

The absence of contamination of fibroblasts and keratinocytes by mycoplasma was confirmed by a Myco Real-Time PCR kit (Evrogen, Moscow, Russia) following the manufacturer’s instructions.

Immortalization of primary fibroblasts and keratinocytes was performed by lentiviral transduction (hTERT & bmi1) according to the previously published protocol (Evtushenko et al.)
